# Deconvolution-based cell-type specific DNA methylation-wide and transcriptome-wide association studies identify risk CpG sites and genes associated with colorectal cancer risk

**DOI:** 10.64898/2026.06.11.26355460

**Authors:** Qing Li, Lili Xu, Jifeng Wang, Chao Li, Wanqing Wen, Xiang Shu, Yaohua Yang, Xiao-ou Shu, Qiuyin Cai, Jirong Long, Bhuminder Singh, Ken S Lau, Zhijun Yin, Graham Casey, Mingyang Song, Ulrike Peters, Wei Zheng, Xingyi Guo

**Affiliations:** Division of Epidemiology, Department of Medicine, Vanderbilt Epidemiology Center, Vanderbilt-Ingram Cancer Center, Vanderbilt University Medical Center, Nashville, TN, USA; Department of Epidemiology and Biostatistics, Memorial Sloan Kettering Cancer Center, New York, NY, USA; Department of Genome Sciences, UVA Comprehensive Cancer Center, School of Medicine, University of Virginia, Charlottesville, VA, USA; Epithelial Biology Center and Department of Cell and Developmental Biology, Vanderbilt University Medical Center, Nashville, TN, USA; Department of Biomedical Informatics, Vanderbilt University Medical Center, Nashville, TN, USA; Departments of Epidemiology and Nutrition, Harvard TH Chan School of Public Health, Boston, MA, USA; Division of Public Health Sciences, Fred Hutchinson Cancer Center, Seattle, WA, USA; Department of Epidemiology, School of Public Health, University of Washington Seattle, WA, USA

**Keywords:** Colorectal cancer, ctMWAS, ctTWAS, Cell-type specific analysis, Susceptibility genes, CpG methylation, *SF3A3*

## Abstract

Bulk tissue-based DNA methylation-wide (MWAS) and transcriptome-wide association studies (TWAS) have identified CpG sites and genes associated with colorectal cancer (CRC) risk, but do not account for cellular heterogeneity. To address this, we developed a deconvolution-informed framework to infer cell-type specific DNA methylation and gene expression profiles from bulk normal colon tissues using reference single-cell epigenomic and transcriptomic datasets. We performed cell-type specific MWAS (ctMWAS) using deconvoluted DNA methylation data from 293 normal colon samples and conducted cell-type specific TWAS (ctTWAS) using deconvoluted gene expression data from 707 normal colon samples. Genetically predicted methylation and expression models were integrated with CRC GWAS summary statistics (78,473 cases and 107,143 controls) to identify risk-associated CpG sites and genes. Through ctMWAS, ctTWAS, and colocalization analyses, we identified 178 high-confidence cell-type-specific CpG sites and 68 genes associated with CRC across 26 previously unreported GWAS loci. Through additional integrative methylation-gene analysis, we prioritized 132 candidate risk genes, the majority of which were supported by multi-omics evidence and stage-specific dysregulation across the adenoma-carcinoma and serrated-carcinoma progression pathways. Pathway enrichment analyses implicated pathways involved in DNA double-strand break repair, TP53 regulation, TGF-β signaling, and innate immune responses. Among prioritized genes, 14 were identified as putative druggable targets linked to 90 FDA-approved or clinical-stage drugs. Experimental validation supports an oncogenic role for *SF3A3*. These findings demonstrate that deconvolution-informed integrative analyses enable cell-type-resolved identification of epigenetic and transcriptional mechanisms underlying CRC susceptibility and provide insights into disease biology, prevention, and therapeutic target discovery.

**Significance:** We developed a deconvolution-informed, cell-type specific multi-omics framework that identified CRC risk-associated CpG sites and susceptibility genes, revealing cell-type specific regulatory mechanisms, key biological pathways, and druggable targets relevant to CRC prevention, therapeutic development, and drug repurposing. Functional validation further supports *SF3A3* as a potential oncogenic driver in colorectal carcinogenesis.

## Introduction

Genome-wide association studies (GWAS) have identified approximately 250 of risk loci associated with CRC susceptibility (1–3). Increasing evidence suggests that epigenetic mechanisms, particularly DNA methylation (DNAm) at cytosine–phosphate–guanine (CpG) dinucleotides, play a central role in mediating the regulatory effects of genetic risk variants on disease susceptibility. DNAm, a key epigenetic mechanism regulating gene expression, chromatin structure, genome stability, is frequently dysregulated in cancer (4,5) and plays a central role in colorectal tumorigenesis (6,7). Integrative multi-omics analyses combining GWAS, DNAm, and transcriptomic data, through approaches such as DNAm or expression quantitative trait locus (mQTL/eQTL), together with advances in methylation-wide association studies (MWAS), and transcriptome-wide association studies (TWAS), as demonstrated in our prior work, have identified thousands of putative risk CpG sites (7–9), and hundreds of putative risk genes (1–3,10–15), providing important insights into functional regulatory mechanisms underlying CRC susceptibility.

Despite these advances, a major limitation of conventional bulk-tissue based MWAS and TWAS is their reliance on bulk tissue data, which fails to account for cellular heterogeneity arising from mixtures of epithelial cells within colon tissues. This cellular heterogeneity can obscure cell-type specific regulatory signals and reduce statistical power to detect biologically relevant associations. Consequently, delineating cell-type specific epigenetic and transcriptional mechanisms is critical but remains a major challenge in CRC research. Recent advances in single-cell epigenomic and transcriptomic technologies, together with computational deconvolution approaches, now provide new opportunities to overcome these limitations and enable cell-type resolved omics-based association analyses. Specifically, the EpiSCORE represents a notable framework that enables virtual microdissection of bulk DNAm data to infer cell-type specific methylation profiles in solid tissues. Building on this approach, the EpiSCORE DNAm atlas (16) provides comprehensive reference panels of cell-type specific DNAm across diverse tissues, facilitating inference of cell-type resolved methylation patterns. In parallel, single-cell RNA sequencing (scRNA-seq), combined with tools such as CIBERSORTx (17), enables estimation of cell-type specific gene expression from bulk tissues. These approaches facilitate investigation of how germline genetic variation influences DNAm and gene expression within specific cellular contexts in colon tissues. In addition, conventional MWAS and TWAS frameworks do not incorporate regulatory context, limiting their ability to prioritize regulatory variants for gene expression prediction. To address this, we previously developed a framework integrating transcription factor (TF) binding information and identified key TF cofactor interactions driving CRC susceptibility through susceptible TF-occupied cis-regulatory elements (STF-CREs) (12). By incorporating prior knowledge of regulatory architecture, our sTF-TWAS (18) approach substantially enhances the power and accuracy for identifying disease-associated susceptibility genes.

In this study, we developed deconvolution-informed cell-type specific multi-omics framework to systematically identify CpG sites and genes associated with CRC risk. We deconvolved bulk DNAm data from 293 individuals of European ancestry using the EpiSCORE framework and tensor composition analysis (TCA) to infer cell-type specific DNAm profiles. These data were used to build cell-type specific models of genetically predicted DNAm, which were further integrated with GWAS summary statistics to perform cell-type specific MWAS (ctMWAS). In parallel, we deconvolved bulk gene expression data from 707 individuals of European ancestry using CIBERSORTx and conducted cell-type specific TWAS (ctTWAS). By integrating epigenomic and transcriptomic models predicted from CRC putative TF-occupied regulatory variants with large GWAS through sTF-TWAS (18), we aim to elucidate cell-type specific epigenetic and transcriptional mechanisms underlying CRC susceptibility and enhance the identification of CpG methylation sites, genes, and regulatory pathways associated with CRC risk.

## Materials and Methods

### Data resources

Bulk DNAm data generated from normal colorectal tissues were obtained from COLONOMICS (n = 132, including 95 normal colon and 37 mucosa) (7,19), and from GTEx (n = 161) (20,21). Bulk gene expression data generated from normal colorectal tissues from BarcUVa-Seq (n = 423) (11,12) and GTEx (n = 284) (20,21). Single-cell RNA-seq data from normal colon tissue tissues (∼30,000 cells, ∼35,000 genes, n = 31) which was utilized to generate single-cell reference matrix was obtained from the Colorectal Molecular Atlas Project (COLON MAP) (22). A reference single-cell DNAm matrix summarizing methylation across 127 marker genes for the colon epithelial cell types was obtained from the EpiSCORE DNA atlas (16). European ancestry CRC GWAS summary statistics (78,473 cases and 107,143 controls) released from previous work (1), was used for ctMWAS and ctTWAS analyses. Additional scRNA-seq data from the COLON MAP (22), included serrated polyps (n = 19), conventional adenomas (n = 29), and carcinomas, including microsatellite-stable (MSS, n = 17) and microsatellite instability-high (MSI-H, n = 15) tumors. These data were used for differential expression analyses to evaluate stage-specific dysregulation along adenoma-carcinoma and serrated-carcinoma sequences. Details of additional datasets, including The Cancer Genome Atlas (TCGA)(23), Clinical Proteomic Tumor Analysis Consortium (CPTAC) (24), DepMap data (25), DrugBank (26), ChEMBL (27), the Therapeutic Target Database(28), and Open Targets(29) were provided in the Data Availability section.

### Construction of genetically predicted cell-type-specific DNAm and gene expression models

Cell-type-specific DNAm and gene expression profiles were inferred from bulk colon tissue datasets using deconvolution-based approaches informed by single-cell references. For DNAm, bulk tissue profiles from the COLONOMICS project (Illumina 450K array, n=132) (7) and the GTEx project (Illumina EPIC array, n=161) (20,21) were processed using standard quality control procedures, including removal of low-confidence probes, probes with ambiguous mapping, and those affected by nearby SNPs (**Supplementary Methods**). Epithelial cell-type proportions were estimated using the EpiSCORE framework (30), focusing on major epithelial populations (enteroendocrine, enterocyte, goblet, paneth-like, and progenitor cells). Bulk methylation profiles were then deconvoluted using TCA (31) to infer CpG-level, cell-type specific methylation values.(**Supplementary Methods**).

For gene expression, bulk RNA-seq data from the BarcUVa-Seq and GTEx projects were processed following the established pipeline (11,12,20,21), including alignment with STAR (32) and quantification using RNA-SeQC (33). The scRNA-seq data from the COLON MAP project were used to generate epithelial cell-type reference signatures after excluding immune and other non-epithelial or low-quality cells and applied quality control filters to retain cells meeting the following criteria: nUMI > 500, nGene > 250, log10GenesPerUMI > 0.8, and mitoRatio < 0.2 (**Supplementary Methods**). Cell-type proportions were estimated using CIBERSORTx (17), and bulk expression profiles were subsequently deconvoluted to obtain cell-type specific expression using the “Impute Cell Expression” module.

Genotype data for the COLONOMICS, BarcUVa-Seq and GTEx cohorts was processed as previously described (11,12) and variants with minor allele frequency (MAF) less than 0.01 (MAF < 0.01), or significant deviations from Hardy-Weinberg equilibrium (*P* < 10^−6^) were excluded (**Supplementary Methods**). In both ctMWAS and ctTWAS, we utilized CRC putative TF-occupied regulatory variants with large GWAS through our previously developed sTF-TWAS approach (18), variants located within 1 Megabase (Mb) of 51 CRC-associated transcription factors (TF) (12). To construct prediction models, DNA methylation (or gene expression) levels were first were first quantile-normalized and inverse normal transformed, and subsequently residualized to remove the effects of population structure and potential confounders, including sex, age, the top five genetic principal components (PCs), and latent factors inferred using Probabilistic Estimation of Expression Residuals (PEER)(34). Following a previous study (21) and the GTEx covariate selection pipeline (20), we applied 20 PEER factors for DNA methylation datasets, 60 for BarcUVa-Seq colon tissue RNA-seq dataset and 45 for the GTEx transverse colon RNA-seq dataset (included factors based on the sample size). The DNA methylation (or gene expression) residuals were inverse normal transformed and utilized in elastic net regression models to estimate the effects of TF-occupied regulatory variants (Equation 1), thereby generating genetically predicted DNA methylation (or gene expression) models for downstream association analyses.

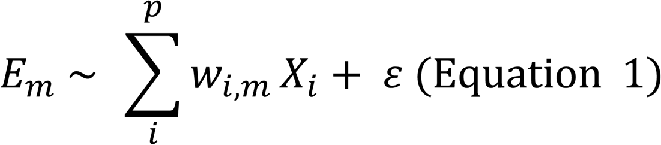

where *E*_*m*_ represents for the *m*-th DNAm level (or gene expression); *X*_*i*_ denotes the genotype vector of the *i*-th variant (*X*_*i*_=0, 1, or 2) and *p* is the total count of variants within the cis-region. The term *w*_*i*,*m*_ represents effect size of *i-th* variant for *m-th* DNAm (or gene expression), and ε is the residual with variance 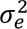. An elastic-net penalty, combining *L*_1_ and *L*_2_ regularizers, is applied to optimize (*w*_*i*,*m*_) for genetic variants. To optimize model parameters and evaluate predictive efficacy, we used ten-fold cross-validation. Model performance was evaluated by calculating the Pearson correlation coefficient (*r*) between the predicted and observed DNAm (gene expression) levels. Models with positive predictive performance (*r* > 0.1) were retained for downstream association analyses.

### Association analysis to identify CpG sites and genes associated with CRC risk

For ctMWAS, we conducted association analyses by integrating genetically predicted CpG models with CRC GWAS summary statistics from 185,616 European individuals using the S-PrediXcan (Equation 2) (35).

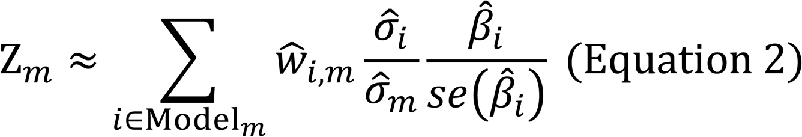

where, the Z_*m*_ estimates the association between predicted DNAm level and CRC risk; 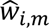 is the weight of genetic variant *i*-th for predicting the level of *g*-th DNAm at CpG site. 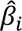 and se (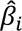) are the GWAS-reported regression coefficients and its standard error for *i*-th variant; and 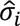 and 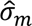 are the estimated variances of *i*-th variant and the predicted *m*-th DNAm level. To combine ctMWAS results from the COLONOMICS and GTEx datasets, we performed a meta-analysis using the aggregated Cauchy association test (ACAT)(36). CpG sites were considered significant at a Bonferroni-corrected threshold of *P* < 0.05, adjusted for the number of tests in each cell type (Enteroendocrine: 193,615; Enterocyte: 176,682; Goblet: 163,721; Progenitor: 166,538).

For ctTWAS, we integrated genetically predicted gene expression models with CRC GWAS summary statistics using S-PrediXcan (35), following the same framework as ctMWAS. Association results from the BarcUVa-Seq and GTEx datasets were subsequently combined through meta-analysis (36). Statistical significance was defined using a Bonferroni-corrected threshold, accounting for the number of genes tested in each cell type (6,856 Absorptive cells, 6,426 Goblet, and 1,104 Stem cell genes).

### Putative risk genes supported by multi-omics and functional analyses

To evaluate functional evidence for putative risk genes, we integrated multi-omics and functional genomic datasets spanning bulk gene expression, proteomic, and single-cell transcriptome data, and CRISPR screening resource. Significant differential gene and protein expression between normal colon and carcinoma tissues were identified using the UALCAN (35,37), incorporating data from TCGA (23) and CPTAC (24) (Welch’s t-test, nominal *P* < 0.05). Based on ctTWAS results, genes for which decreased expression was associated with increased CRC risk were classified as putative tumor suppressors, whereas genes for which increased expression was associated with elevated risk were classified as putative oncogenes. Accordingly, tumor suppressor candidates were considered functionally supported if they exhibited significantly lower expression in carcinoma compared to normal tissue, with the opposite pattern expected for putative oncogenes. A similar framework was applied to evaluate putative ctMWAS-associated genes (**Supplementary Methods**). Gene essentiality was assessed using CRISPR–Cas9 screening data from the Cancer Dependency Map (25) (DepMap, 26Q1) based on 64 colorectal adenocarcinoma cell lines, with genes exhibiting median CERES scores less than −0.5 across cell lines defined as essential for cell proliferation and survival.

To further evaluate functional relevance along disease progression, we analyzed scRNA-seq data from the COLON MAP project (38), encompassing serrated polyps, conventional adenomas, and colorectal carcinomas. Pseudobulk gene expression profiles were constructed for cell-type specific populations, and differential expression analyses were performed using DESeq2 (39) to compare adjacent stages along the adenoma–carcinoma and serrated-carcinoma etiological pathways. Specifically, we focused on two major colorectal carcinogenesis sequences, adenoma–carcinoma and serrated polyp–carcinoma. We compared gene expression profiles across single-cell populations, including adenoma-specific cells (ASC) or serrated-specific cells (SSC) and carcinoma cell populations, including microsatellite-stable (MSS and microsatellite instability–high (MSI-H) tumor subtypes. Differentially expressed putative risk genes with statistically significant changes false discovery rate (FDR < 0.05) were considered to have functional evidence supporting their role in CRC progression.

### Cell-type specific pathway enrichment analysis

To characterize the biological functions and pathways associated with cell-type specific genes, we performed enrichment analysis for CRC risk genes identified from each cell type using Enrichr (40). Enrichr integrates multiple curated databases for comprehensive functional annotation. Enrichment significance was assessed using a combined score that incorporates the Fisher’s exact test *P* value and a Z score reflecting deviation from the expected rank. Pathways with FDR < 0.05 were considered significantly enriched.

### siRNA knockdown assay

Human colon cancer cell lines SW480, RKO and HCT116 were obtained from the American Type Culture Collection (ATCC, Manassas, VA). They were cultured in RPMI 1640 supplemented with 2 mM L-glutamine, 100 IU/mL penicillin-streptomycin, 1 mM sodium pyruvate, 10 mM HEPES, 1.5 mg/L sodium bicarbonate, and 10% fetal bovine serum (FBS), at 37°C in a humidified atmosphere with 5% CO₂. Total RNA was isolated from the three colon cancer cell lines using the miRNeasy Mini Kit (Qiagen) according to the manufacturer’s protocol. Complementary DNA (cDNA) was synthesized using the High-Capacity cDNA Reverse Transcription Kit (Thermo Fisher Scientific). qPCR amplification was performed on a CFX384 Touch Real-Time PCR Detection System (Bio-Rad) using Luna Universal qPCR Master Mix (New England BioLabs). Relative mRNA expression levels were calculated using the ΔΔCt method. *SF3A3* gene expression (*SF3A3* probe, Assay ID:ID: Hs00757649_s1, Thermo Fisher Scientific Inc) was normalized to glyceraldehyde-3-phosphate dehydrogenase (GAPDH probe, Assay ID: Hs02786624_g1) as an internal control. Relative transcript levels were calculated using the comparative cycle threshold (Ct) method. Additional details on cell viability, migration, invasion, and colony formation assays are provided in the **Supplementary Methods**.

## Results

### Overview of study design and analytic framework

To resolve cell-type specific mechanisms underlying CRC susceptibility, we developed an integrative analytical framework that deconvolves bulk DNAm and gene expression data into cell-type resolved profiles, enabling ctMWAS and ctTWAS (**Figure 1**). We analyzed bulk tissues-based DNAm from 293 (n=132 from COLONOMICS and n=161 from GTEx) normal colon samples of European ancestry. After quality control and normalization (**Materials and Methods**), 430,086 and 754,119 CpG probes were retained in the COLONOMICS and GTEx datasets, respectively. After deconvoluting bulk tissue DNAm profiles using the EpiSCORE framework (**Materials and Methods**), our results showed consistent epithelial cell-type composition across both cohorts, with enteroendocrine cells representing the largest fraction, followed by progenitor, goblet, and enterocyte cells (**Tables S1-S2**). Paneth-like cells exhibited consistently low abundance and were excluded from downstream analyses. We then applied TCA to infer cell-type specific methylation levels for each CpG site across samples, generating imputed DNAm profiles for the four major epithelial cell types. Using the deconvoluted DNAm profiles, we conducted ctMWAS studies by applying genetically predicted DNAm to CRC GWAS summary statistics from European ancestry to identify cell-type specific CpG sites associated with CRC risk (**Figure S1**).

**Figure 1:**
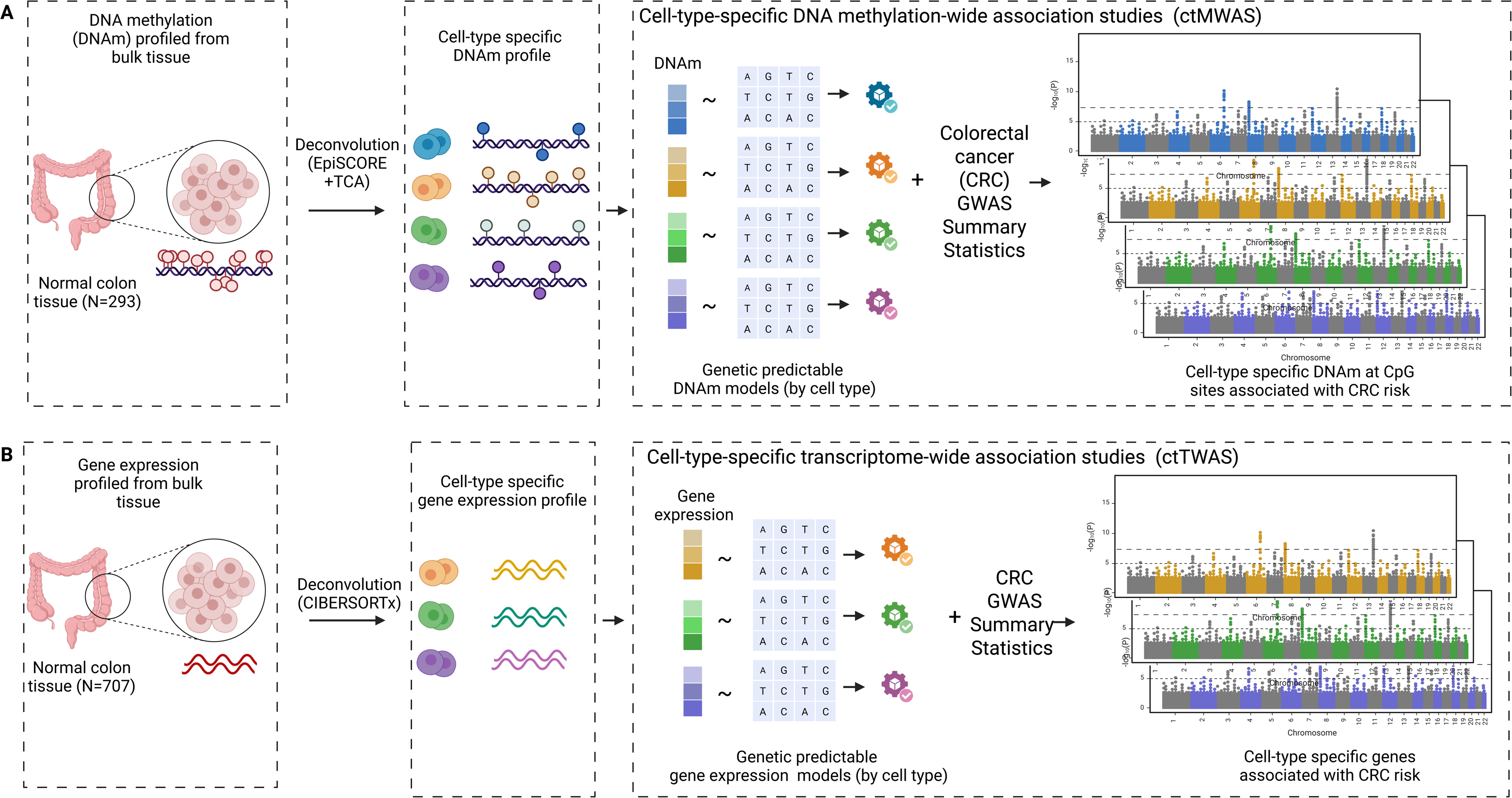
Overview of the analytical framework. Schematic illustration of the integrative pipeline for identifying cell-type specific DNAm CpGs sites and gene expression associated with colorectal cancer risk. **A)** Bulk DNA methylation data from normal colon tissue were deconvoluted into cell-type specific methylation profiles using EpiSCORE and tensor composition analysis (TCA), guided by single-cell reference methylation data. Cell-type specific methylation prediction models were then integrated with CRC GWAS summary statistics to perform ctMWAS and identify risk-associated CpG sites. **B)** Bulk RNA-seq data from xx datasets were deconvoluted into cell-type specific gene expression profiles using CIBERSORTx with single-cell reference expression data. Cell-type specific gene expression prediction models were then integrated with CRC GWAS summary statistics to perform ctTWAS and identify risk-associated genes.

In parallel, we analyzed bulk gene expression data from 707 individuals of European ancestry (n=423 from BarcUVa-Seq and n=284 from GTEx). Using the reference scRNA-seq in normal colon tissues from the COLON MAP (22) and CIBERSORTx tool(17), we estimated epithelial cell-type composition across samples (**Materials and Methods**). Our results showed that absorptive cells (enterocytes) constitute the largest fraction, followed by stem (progenitor) and goblet cells, consistent with the DNAm based estimates (**Tables S3-S4**). We next inferred their cell-type specific gene expression profiles for all samples using the Impute Cell Expression function in CIBERSORTx. Using the deconvoluted expression profiles, we performed ctTWAS for the three major epithelial cells through constructing genetic predictive models of gene expression and integrating these models with CRC GWAS summary statistics from European ancestry to identify cell-type specific genes associated with CRC risk (**Figure S2**).

### Identification of cell-type-specific CpG sites associated with CRC risk

We conducted ctMWAS based on deconvoluted DNAm profiles for each epithelial cell type in both COLONOMICS and GTEx datasets (**Materials and Methods**). In the COLONOMICS dataset, 64,355, 60,569, 58,076 and 63,280 genetically predicted methylation models were built for enteroendocrine, enterocyte, goblet and progenitor cells, respectively (*r* > 0.1, **Materials and Methods**). In the GTEx dataset, 144,349, 125,804, 113,808 and 112,037 predictable models were built for the corresponding cell types. We then performed ctMWAS separately within each dataset and combined the results through meta-analysis of summary statistics across datasets (**Materials and Methods**). At a Bonferroni-corrected *P* < 0.05, we identified 2,341 significant cell-type-specific CpG–CRC risk associations, representing 2,134 unique CpG sites (**Figure 2, Table S5**). These associations included 681 in enteroendocrine cells, 496 in enterocytes, 726 in goblet cells, and 438 in progenitor cells and mapped to 249 independent loci (73, 69, 59, and 48 loci, respectively). To prioritize likely causal signals, we performed colocalization analyses between cell-type-specific mQTLs and CRC GWAS (**Materials and Methods**). We identified 178 high-confidence CpG sites in 106 loci (43% of all loci) with strong evidence for a shared causal variant between DNA methylation and CRC risk (PP.H4 > 0.8), including 37 loci in enteroendocrine cells, 30 in enterocytes, 20 in goblet cells, and 19 in progenitor cells (**Table S5**). Notably, 14 of these loci were novel CRC susceptibility loci, including six in enteroendocrine cells, five in enterocytes, one in goblet cells, and two in progenitor cells, each located more than 1 Mb from previously reported CRC GWAS lead variants (2). To further assess whether ctMWAS-identified CpG sites influence CRC susceptibility through cis-regulation of gene expression, we additionally examined associations between 178 high-confidence CpGs and nearby genes (±1 Mb) using matched DNAm and gene expression data from the GTEx samples (**Materials and Methods**). We identified 108 CpG–gene pairs with significant negative correlations (nominal *P* < 0.05), corresponding to 86 unique genes potentially regulated by these methylation sites (**Table S6**).

**Figure 2:**
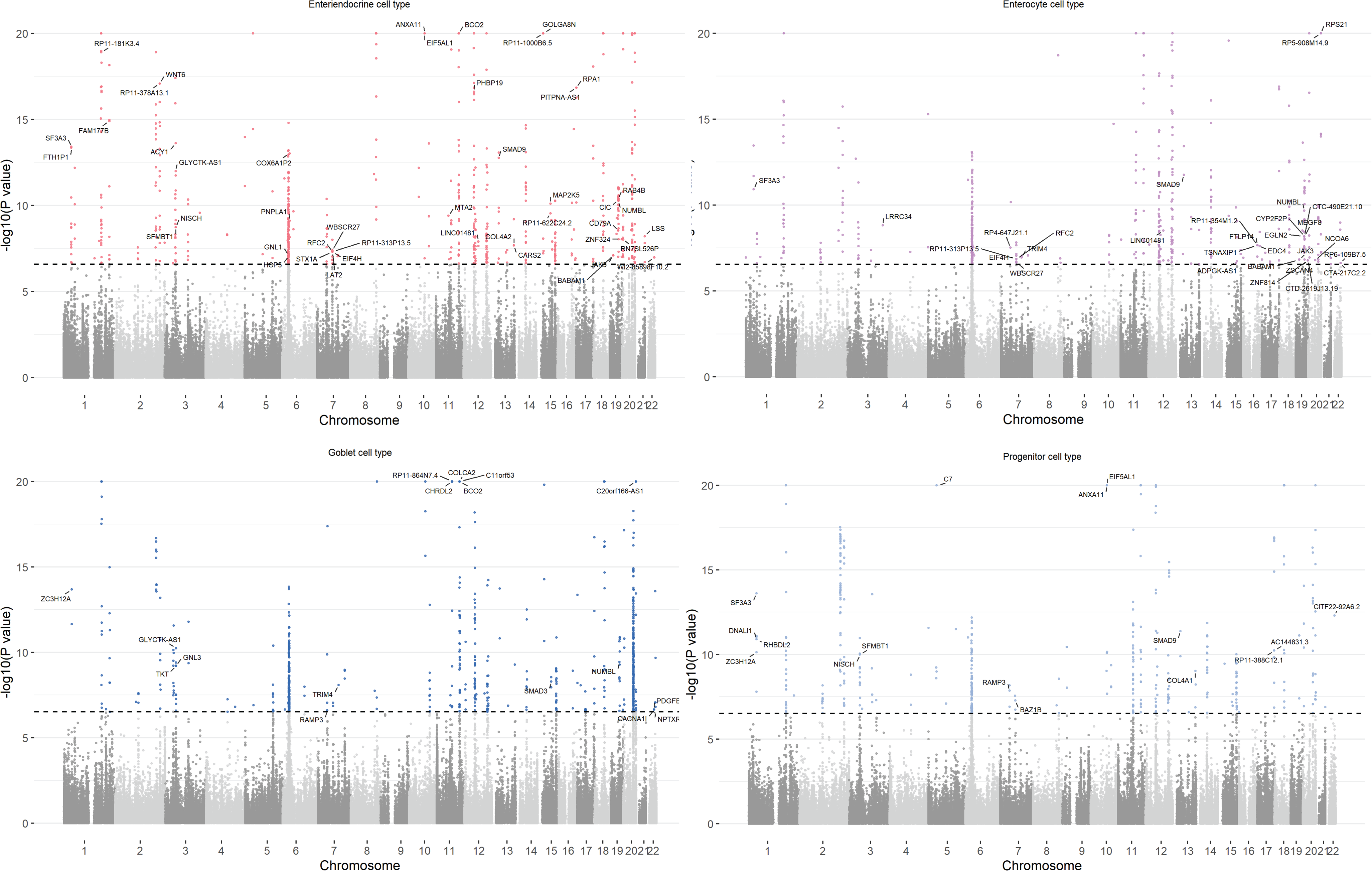
Cell-type specific DNA methylation associated with CRC risk. Manhattan plots of ctMWAS results across epithelial cell types. Each point represents a CpG site, and dashed lines indicate Bonferroni-corrected significance thresholds. Panels correspond to **A)** enteroendocrine, **B)** enterocyte, **C)** goblet, and **D)** progenitor cells. High confidence CpGs site (PP.H4 > 0.8) are labeled with their corresponding target genes.

### Identification of cell-type specific genes associated with CRC risk

We conducted ctTWAS based on the deconvoluted gene expression profiles for each epithelial cell (absorptive, goblet, and stem cell types) from 707 participants of European ancestry (**Materials and Methods**). In the BarcUVa-Seq dataset, 5921, 641 and 3900 genetically predicted expression models were built for absorptive, goblet, and stem cell types, respectively. In the GTEx dataset, 2127, 492 and 3967 predicted models were constructed for the corresponding cell types. We then performed ctTWAS separately within each dataset and combined the results through meta-analysis of summary statistics across datasets (**Materials and Methods**). At a Bonferroni-corrected *P* < 0.05, we identified 151 significant cell-type specific gene-CRC risk associations (representing 121 unique genes), including 84 associations in absorptive cells, 61 in stem cells, 6 in goblet cells (**Figure 3, Table S7**). Further colocalization analyses (**Materials and Methods**) identified 68 high-confidence gene–CRC risk associations, corresponding to 53 unique genes, with strong evidence for a shared causal variant between gene expression and CRC risk (PP.H4 > 0.8). The high-confidence associations included 39 genes in absorptive cells, 26 in stem cells, and 3 in goblet cells (**Table S7**). Notably, 12 of these associations mapped to novel CRC susceptibility loci, including five in absorptive cells and seven in stem cells.

**Figure 3:**
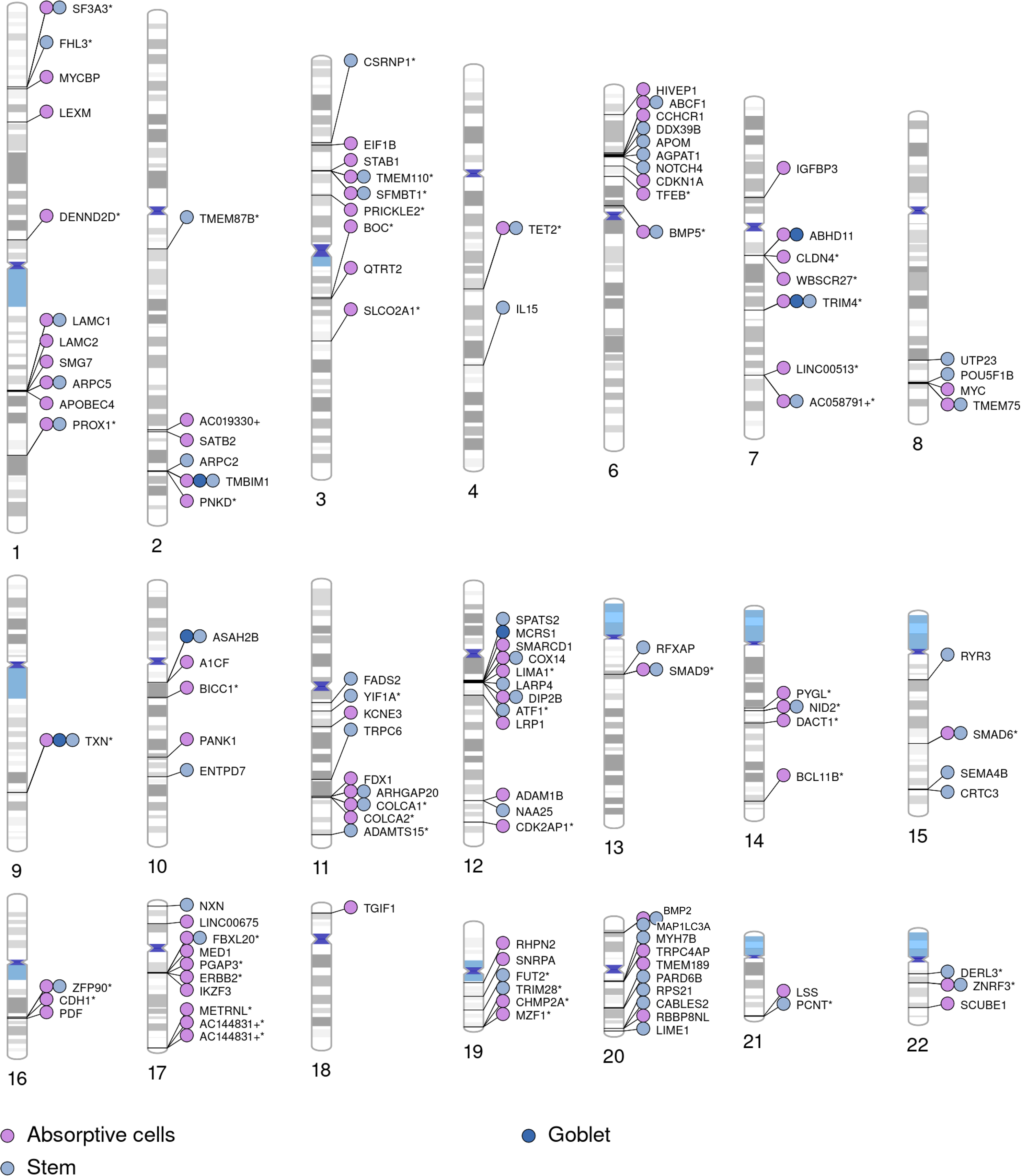
Cell-type-specific genes associated with colorectal cancer (CRC) risk. Distribution of ctTWAS-identified genes across epithelial cell types. Genes are colored according to their corresponding cell type. An asterisk (*) denotes genes with strong colocalization support (PP.H4 > 0.8). A plus sign (+) indicates that the gene name was truncated due to space limitations in the figure.

### Functional characterization of cell-type specific CRC risk genes

To functionally evaluate risk genes identified by ctMWAS and ctTWAS, we analyzed 132 unique high-confidence CRC susceptibility genes, comprising 86 methylation-associated genes from ctMWAS and 53 genes from ctTWAS, with 7 genes shared between the two approaches (**Materials and Methods**). We evaluated their functional relevance using complementary multi-omics evidence from independent transcriptomic, proteomic, and functional genomic datasets. At the transcriptomic level, 34 genes (11 ctMWAS and 25 ctTWAS) showed differential expression between normal colon and carcinoma tissues in The Cancer Genome Atlas (TCGA)(23) (nominal *P* < 0.05), with effect directions concordant with ctMWAS/ctTWAS-inferred functional roles (e.g., oncogenic or tumor-suppressive) (**Figure 4A-B, Table S8**). Consistent evidence was observed at the proteomic level, where 12 proteins (5 ctMWAS and 8 ctTWAS) showed differential abundance in Clinical Proteomic Tumor Analysis Consortium (CPTAC) data (24) (nominal *P* < 0.05) (**Figure 4C-D, Table S8**). In addition, functional genomic screening using DepMap data (25) identified 5 genes (3 ctMWAS and 3 ctTWAS) as essential for CRC cell viability (**Figure 4E-F, Table S8**).

**Figure 4.**
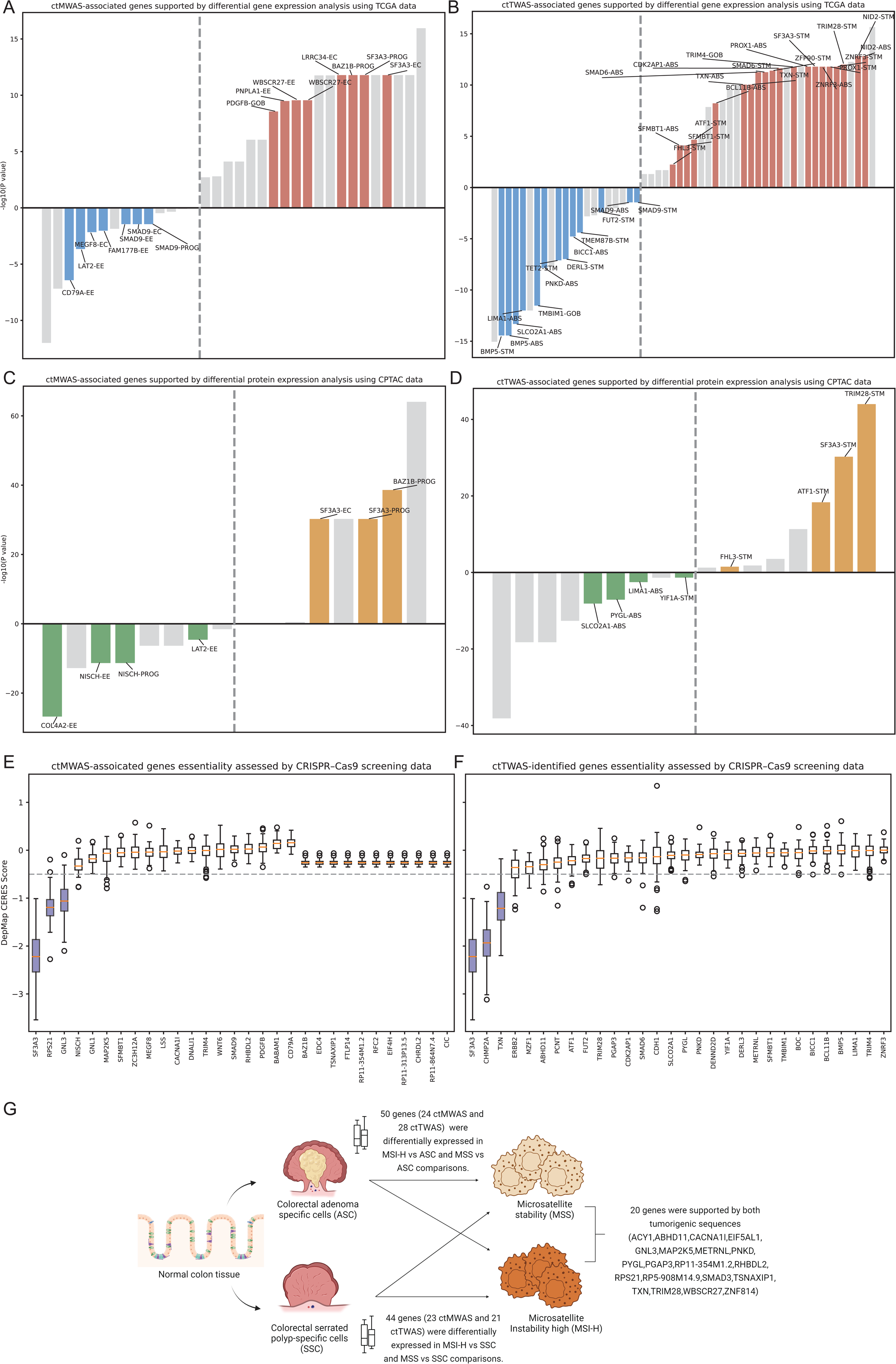
Multi-omics and single-cell functional support for prioritized CRC risk genes. Functional validation of ctMWAS- and ctTWAS-prioritized genes using multiple data sources. TCGA-supported genes showing differential gene expression (*P* < 0.05) with concordant directionality (up- or downregulation) for **(A)** ctMWAS and **(B)** ctTWAS. CPTAC-supported genes showing differential protein expression (*P* < 0.05) with concordant directionality for **(C)** ctMWAS and **(D)** ctTWAS. Genes essential for cell proliferation identified from DepMap CRISPR–Cas9 screening for **(E)** ctMWAS and **(F)** ctTWAS. Boxes are colored purple when the median essentiality score falls below 0.5, indicating gene essentiality, and left unfilled otherwise. **(G)** Single-cell differential expression across CRC progression, including the adenoma–carcinoma and serrated polyp–carcinoma sequences.

To further characterize their roles along colorectal tumorigenesis, we performed single-cell differential expression analysis using scRNA-seq data focusing on the two major CRC sequences, adenoma–carcinoma and serrated polyp–carcinoma (**Materials and Methods**). Along the adenoma–carcinoma sequence, 50 genes (24 ctMWAS and 28 ctTWAS) were differentially expressed (FDR < 0.05), with effect directions of concordant with ctMWAS/ctTWAS results(**Figure 4G, Table S9**). Along the serrated polyp–carcinoma sequence, 44 genes (23 ctMWAS and 21 ctTWAS) showed concordant effect directions of concordant with ctMWAS/ctTWAS results (**Figure 4G, Table S9**). Notably, 20 genes were supported by both tumorigenic sequences, suggesting the presence of shared core susceptibility genes and biological mechanisms that may play fundamental roles in colorectal cancer initiation and progression (**Table S9**).

### Cell-type specific pathway enrichment

To elucidate the biological mechanisms underlying CRC susceptibility, we performed pathway enrichment analyses of ctMWAS- and ctTWAS-prioritized genes using Enrichr including Reactome, MSigDB Hallmark, and KEGG databases (**Materials and Methods**). For the ctMWAS-derived genes, 33 significant enriched pathways were shown in Enteroendocrine cells models (False Discovery Rate < 0.05) (**Table S10**). Among them, multiple pathways from homologous recombination repair (HRR; e.g., Homology Directed Repair, Presynaptic Phase of Homologous DNA Pairing and Strand Exchange, Defective HRR Due to *BRCA2* Loss of Function) indicate that impaired HRR is a pervasive feature of CRC, consistent with somatic *BRCA2* dysfunction and *RAD51* mediated repair deficiency driving genomic instability. In addition, DNA double-strand break repair pathways (DSBR; including Diseases of DNA Double-Strand Break Repair and Processing of DNA Double-Strand Break Ends) further implicate DSBR failure as a key contributor to the chromosomal instability characteristic of CRC. Additionally, enrichment of DNA damage checkpoint and TP53 regulation pathways (G2/M DNA Damage Checkpoint, Regulation of TP53 Activity) reflects the well-established role of checkpoint abrogation in CRC progression, consistent with *TP53* being mutated in approximately 60% of non-hypermutated CRC cases (41).

For ctTWAS-derived genes, pathway enrichment was largely confined to absorptive cells (4 pathways), goblet (36 pathways) and one pathway for stem cells (**Table S10**). In absorptive and stem cells, enrichment centered on TGF-β signaling. In goblet cells, enriched pathways followed a sequential progression from inflammatory signaling through oxidative stress response to transcriptional reprogramming. Innate immune pathways were broadly activated, including NF-κB signaling via *TRAF6* and *FADD/RIP-1, NLRP3* inflammasome assembly, and IRF-mediated interferon induction, all of which contribute to the chronic mucosal inflammatory environment that drives the adenoma–carcinoma sequence. This sustained inflammation was accompanied by enrichment of ROS detoxification pathways and the *KEAP1–NFE2L2* antioxidant axis, indicating that pre-neoplastic epithelial cells experience both oxidative damage and compensatory stress protection simultaneously. Further downstream, reprogramming of *FOXO, TP53,* and *NFE2L2* transcriptional networks suggests that these key regulators shift their activity in response to oxidative injury, facilitating the transition toward malignant transformation.

### CRC risk genes as potential druggable targets

To assess therapeutic and prevention relevance, we examined the identified risk genes through databases of approved and investigational drug-protein interactions (**Supplementary Methods**). Among the 132 putative CRC risk genes, 14 were identified as putative druggable targets, which are currently targeted by 90 approved or clinical-stage compounds, including 66 inhibitors, eight binders, five activators, and three modulators (**Table S11**). Notably, five genes (*CACNA1I, ERBB2, JAK3, NISCH* and *TXN*) are targeted by 43 drugs currently under clinical evaluation for cancer treatment (**Figure 5),** including three novel genes (*CACNA1I*, *JAK3*, and *NISCH*) identified through ctMWAS. Specifically, our findings provide genetic support for several clinically relevant therapeutic targets that are currently being investigated in CRC or other cancers, including *ERBB2* (e.g., Sapitinib and Pyrotinib), ***JAK3*** (e.g., AT-9283), *TXN* (e.g., PX-12), and *NISCH* (e.g., Tepotinib and Moxonidine). The identification of these clinically actionable targets from our genetic analyses highlights promising opportunities for CRC therapeutic development and drug repurposing.

**Figure 5:**
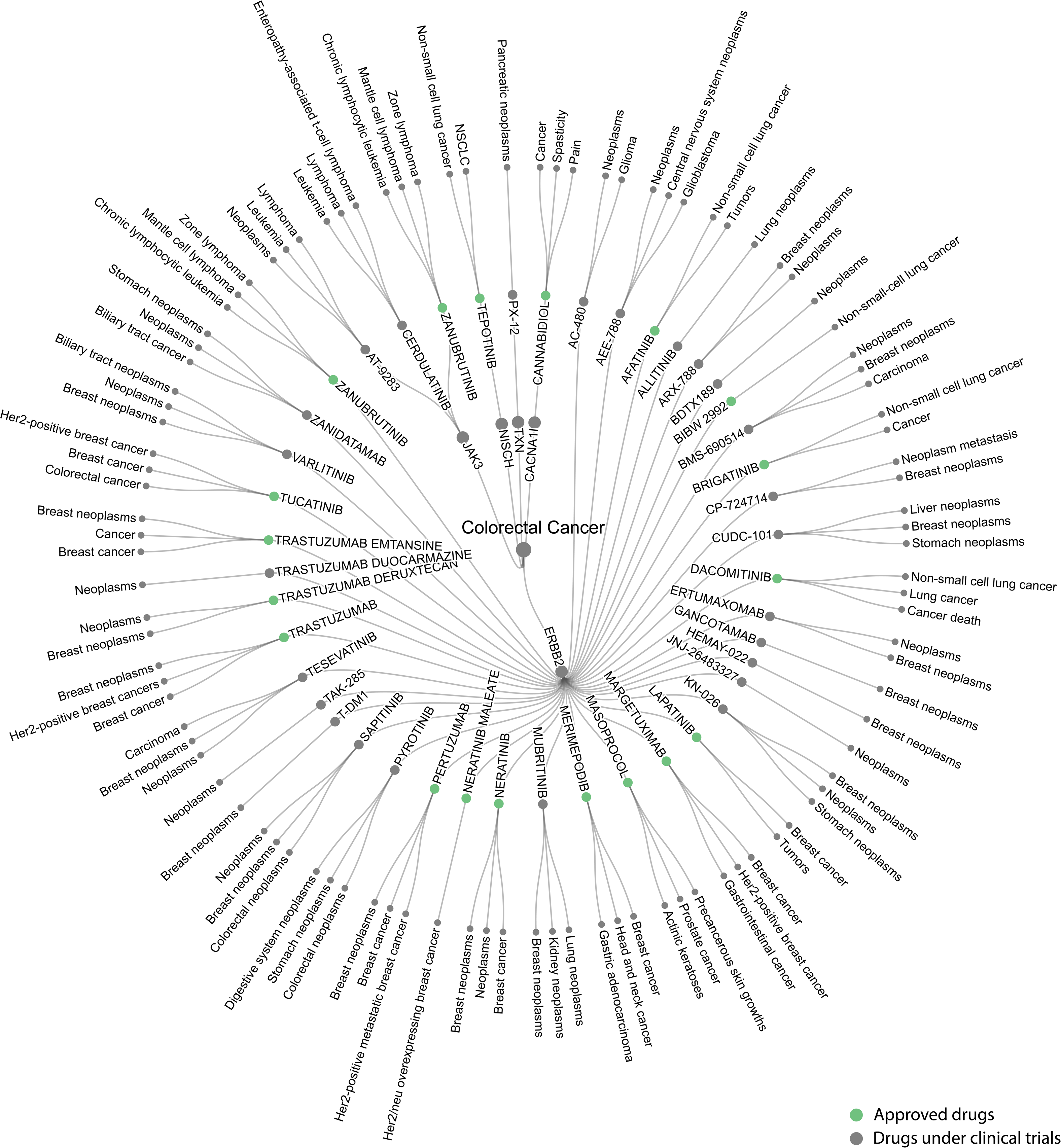
Druggable CRC risk genes and associated cancer therapeutics. From innermost to outermost layers: the inner circle shows 5 risk genes; the middle circle displays 43 drugs targeting these genes, with green circles indicating those approved and grey circles indicating those in clinical trials. The outermost circle shows shortened cancer indications for each drug, with the most relevant indications (up to three) shown per drug.

### Functional assays for *SF3A3* as a putative oncogene

Among the genes commonly identified by both ctMWAS and ctTWAS, we identified six were associated with CRC risk within the same epithelial cell lineage, providing convergent evidence from both methylation- and expression-based analyses for their potential causal roles in colorectal carcinogenesis (**Table S12**). Of note, two genes (*SF3A3 and SMAD9*) exhibited concordant functional roles based on ctMWAS and ctTWAS inference (**Supplementary Methods**). We next prioritized *SF3A3* for functional validation as it showed consistent oncogenic signal across two epithelial cell types, including enterocytes and progenitor cells. siRNA-mediated knockdown of *SF3A3* (**Materials and Methods**) in three CRC cell lines (HCT116, RKO, and SW480) achieved efficient suppression (*P* < 0.05) at the mRNA level, as confirmed by RT–PCR (**Figure 6A**, **Table S13**). *SF3A3* depletion significantly reduced cell viability in HCT116 (*P* = 1.6 × 10^-2^), with moderate reductions observed in RKO and SW480 (**Figure 6B**). Functionally, *SF3A3* knockdown markedly impaired migration and invasion capacities in HCT116 (*P* = 1.88 × 10^-2^ and 4.60 × 10^-2^, respectively) (**Figure 6C-D**), and significantly decreased clonogenic growth across three cell lines (*P* < 0.05) (**Figure 6E-H**). Together, these results provide experimental support for the predicted oncogenic role of *SF3A3* based on our ctMWAS and ctTWAS.

**Figure 6:**
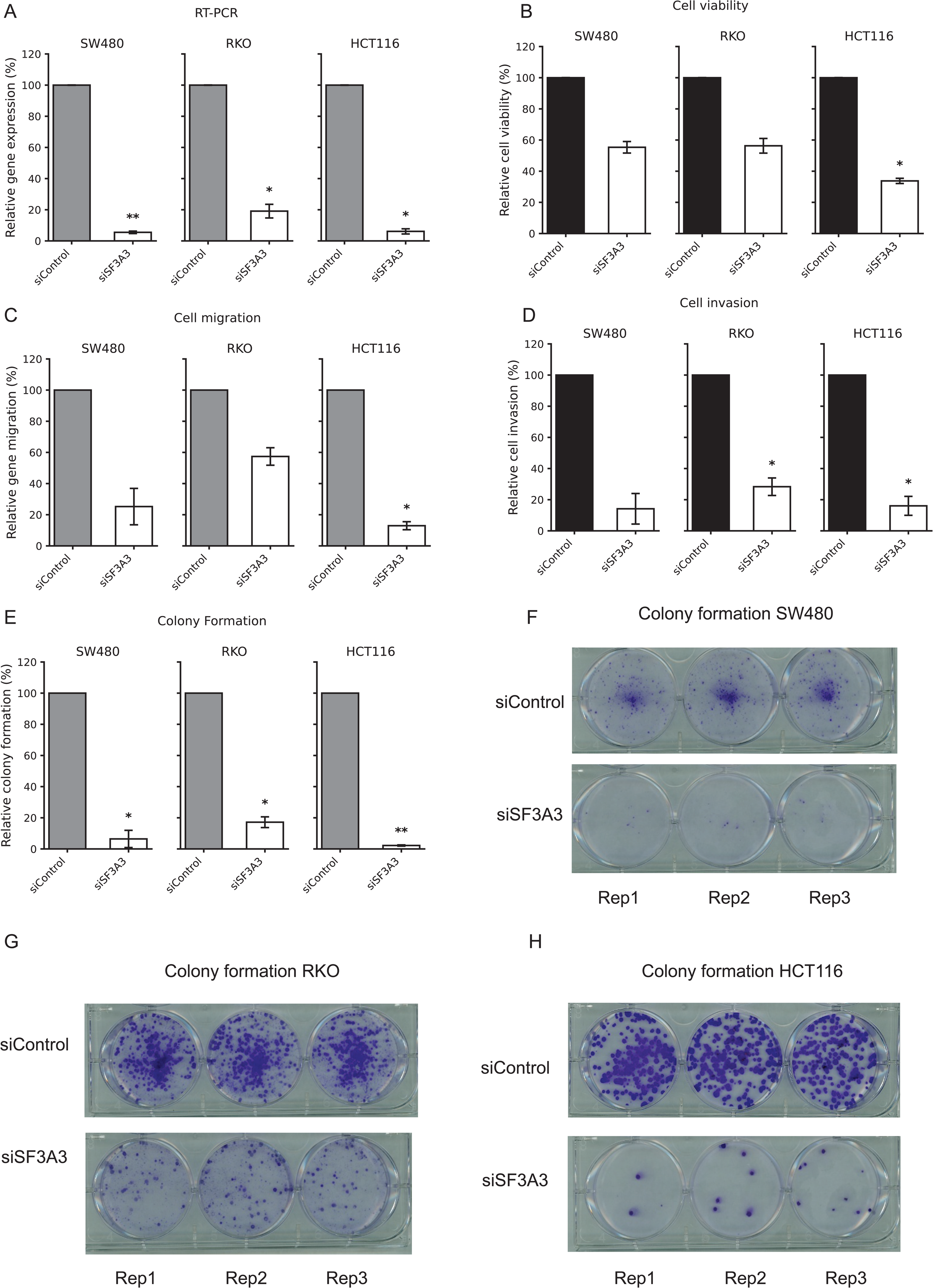
Cell viability and colony formation for the selected gene *SF3A3* in three CRC cell lines (SW480, RKO, HCT116). CRC cell lines with knocking down by one sgRNAs were compared to CRC cell lines with non-targeting sgRNA control. RT-PCR was conducted with miRNeasy Mini Kit. **A)** Cell viability was conducted with Alamar Blue assay **B)** Cell migration **C)** and invasion **D)** were performed in 24-well plates with 8 μm pore-size chamber inserts. Colony formation assays were performed with cell stained by crystal violet (**E-H**). Three independent experiments were performed. P-values were determined by a two-side t-test from the comparison of knockdown and control cells. “*”, *P* < 0.05; “**”, *P* < 0.01.

## Discussion

In this study, we developed an integrative, deconvolution-informed multi-omics framework to delineate cell-type-specific genetic regulatory mechanisms underlying CRC susceptibility. By coupling ctMWAS and ctTWAS with complementary deconvolution algorithms, EpiSCORE/TCA for DNA methylation and CIBERSORTx for gene expression, we systematically linked germline genetic variation to epigenetic and transcriptional regulation within discrete epithelial populations, addressing a major limitation of conventional bulk-tissue analyses. The robustness of this framework is substantiated at multiple levels: each deconvolution algorithm has undergone rigorous independent validation; estimated cellular composition was biologically coherent with the known histological architecture of normal colonic epithelium; and concordance with prior findings provides orthogonal support, with 77% of identified CpGs (1650/2134, **Table S5**) correlating with MWAS signals (|*r*| > 0.4) identified in previous work (7–9) and 83% of identified genes (100/121, **Table S7**) overlapping with prior TWAS discoveries (1–3,10–15). Of note, most previously identified risk genes lack cell-type-specific regulatory characterization. Through integrating cell-type-specific methylation and gene expression analyses, our framework elucidates the cellular contexts and epigenetic regulatory mechanisms underlying both known and newly identified CRC risk genes, providing a more comprehensive and mechanistic understanding of CRC genetic architecture.

Integrating ctMWAS and ctTWAS results, we prioritized 132 high-confidence genes, including 26 unreported CRC-associated GWAS loci and 69 previously unreported CRC-associated genes. The majority of genes were supported by convergent transcriptomic (TCGA), proteomic (CPTAC), and functional genomic (DepMap) evidence, as well as stage-specific dysregulation along the adenoma–carcinoma and serrated–carcinoma sequences. Pathway enrichment analyses revealed distinct cell-type-specific mechanisms: ctMWAS-derived genes were predominantly enriched in homologous recombination repair and DNA double-strand break repair pathways in enteroendocrine cells, whereas ctTWAS-derived genes highlighted TGF-β signaling in absorptive cells and oxidative stress and transcriptional regulation in goblet cells, consistent with epithelial plasticity and tumor progression. Collectively, this framework substantially expands the catalogue of CRC-associated regulatory variation beyond bulk-tissue approaches, offering a scalable alternative to single-cell molecular QTL studies.

A key strength of this study is the integration of ctMWAS and ctTWAS, which provides complementary yet convergent insights into CRC biology. ctMWAS captures genetically regulated DNAm changes that may act through cis-regulatory mechanisms, whereas ctTWAS identifies downstream gene expression effects more directly linked to disease risk. By combining these approaches, we delineate a multi-layered regulatory cascade from genetic variation to epigenetic modification and transcriptional output at a cell-type specific level. Genes supported by both frameworks represent high-confidence candidates with consistent regulatory and functional evidence. For example, *SF3A3* was inferred as oncogenic, as CpGs linked to *SF3A3* exhibited tumor-suppressive patterns in progenitor and enterocyte cells, while ctTWAS supported a concordant oncogenic role in CRC risk in these cell types. This is consistent with prior studies indicating that *SF3A3*, a spliceosome component, promotes tumor growth and RNA splicing dysregulation in breast and colorectal cancer (42,43).. In contrast, a subset of genes showed consistent tumor suppressor roles across both analyses. For example, *SMAD9*, a component of the BMP/TGF-β signaling pathway, is involved in cell differentiation and tumor suppression (44,45). Notably, *TRIM4* exhibits as a tumor suppressor in enterocytes. Mechanistically, *TRIM4* degrades *TPL2* to restrain Wnt/β-catenin signaling, whereas its loss promotes β-catenin activation and tumor progression (46). Consistently, higher *TRIM4* expression has been associated with reduced CRC risk, although evidence from other cancers supports an oncogenic role through promoting proliferation and therapy resistance(47). These results underscore the value of integrative analyses in strengthening causal inference and prioritizing biologically meaningful targets.

Beyond direct therapeutic applications, our findings offer a genetically informed rationale for CRC prevention. The 90 compounds mapped to CRC risk genes include several with established indications in conditions predisposed to CRC, suggesting opportunities for repurposing to interrupt early neoplastic progression. Notably, *JAK3* is targeted by AT-9283, a clinical-stage inhibitor with demonstrated antitumor activity in preclinical CRC models (43) and *TXN* is targeted by PX-12, with demonstrated cytotoxic and anti-migratory effects in CRC cells under hypoxic conditions (44). Together, *JAK3* and *TXN* converge on STAT3-mediated inflammatory signaling, and *NISCH* implicates integrin-driven invasion, pathways with recognized roles in early colorectal neoplasia. Moreover, the prevention rationale is further strengthened by the inflammatory bowel disease (IBD–CRC) link. Tofacitinib, approved for ulcerative colitis, and JAK3-selective inhibitors under investigation for Crohn’s disease target the same *JAK–STAT* axis implicated in our genetic findings. Given that IBD confers a two- to three- increased CRC risk (49,50), *JAK3* inhibition may simultaneously treat IBD and attenuate the inflammatory microenvironment driving colitis-associated CRC. Functional validation in organoid and adenoma models will be essential to bridge these genetic findings with actionable prevention and treatment strategies for CRC.

Despite these strengths, several limitations should be considered. First, although deconvolution methods enable inference of cell-type specific signals, they cannot fully capture the complexity of cellular heterogeneity and may introduce potential estimation error. The lack of matched measured single-cell DNAm data in colon tissue limits direct validation of inferred methylation profiles. Second, our analyses were restricted to individuals of European ancestry, which may limit generalizability; future studies in diverse populations are needed to capture ancestry specific regulatory mechanisms. Third, while we provide strong statistical and multi-omics support, experimental validation remains limited to a subset of genes. Functional studies, including CRISPR-based perturbation and epigenome editing, will be essential to establish causality. Finally, integration of additional single-cell multi-omics data, including chromatin accessibility and methylation, will further refine the regulatory landscape of CRC.

In summary, this study provides a comprehensive framework for identifying cell-type specific regulatory mechanisms underlying CRC susceptibility. By integrating genetic, epigenetic, and transcriptomic data within defined cellular contexts, as well as drug-protein database, we uncover novel risk CpG sites, genes, and CRC-related pathways, and provide additional insights into disease biology as well as druggable targets relevant to CRC prevention, therapeutic development, and drug repurposing. These findings highlight the importance of cell-type resolution in complex disease genetics and offer an opportunity for future functional and translational studies aimed at improving CRC prevention and precision oncology.

## Data availability

Colorectal cancer GWAS summary statistics are publicly available through the **GWAS Catalog** (**GCST90129505**; https://www.ebi.ac.uk/gwas/studies/GCST90129505).

Bulk DNA methylation data from the BarcUVa-Seq project (Illumina 450K arrays) are available via the **COLONOMICS project** (https://www.colonomics.org), and corresponding genotype data can be obtained upon request at EGA (dataset https://ega-archive.org/datasets/EGAD00010001253). Bulk RNA-seq gene expression data from BarcUVa-Seq are accessible through **dbGaP** (**phs003338.v1.p1**; https://www.ncbi.nlm.nih.gov/projects/gap/cgi-bin/study.cgi?study_id=phs003338.v1.p1).

GTEx DNA methylation data are available through the Gene Expression Omnibus (GEO) (**GSE213478**; https://www.ncbi.nlm.nih.gov/geo/query/acc.cgi?acc=GSE213478). GTEx genotype and gene expression data (version 8) are available via **dbGaP** (**phs000424.v11.p2**; https://www.ncbi.nlm.nih.gov/projects/gap/cgi-bin/study.cgi?study_id=phs000424.v11.p2).

Single-cell RNA-seq data from human colon tissue were obtained from the **Colorectal Molecular Atlas Project (COLON MAP)**(38) via the Human Tumor Atlas Network (HTAN) Data Portal (https://humantumoratlas.org/). Cell-type specific DNA methylation reference data were obtained from the **EpiSCORE DNAm atlas** (https://www.biosino.org/episcore).

The publicly available differential gene and protein expression data used in this study were accessed through **UALCAN** (https://ualcan.path.uab.edu/index.html). CRISPR-Cas9 gene essentiality data (DepMap Public 26Q1 CERES scores) were obtained from the **DepMap portal** (https://depmap.org/portal/). Gene annotation data (GENCODE v26, GRCh38) were downloaded from **GENCODE** (https://www.gencodegenes.org/human/release_26.html).

Information on approved cancer drugs was retrieved from the **National Cancer Institute** (https://www.cancer.gov/about-cancer/treatment/drugs). Drug-target and compound data were compiled from multiple publicly available databases, including **ChEMBL** (https://www.ebi.ac.uk/chembl/), the **Therapeutic Target Database (TTD)** (https://db.idrblab.net/ttd/), **Open Targets** (https://www.opentargets.org/), and **DrugBank** (https://go.drugbank.com/).

## Code availability

Analytic code used in this study is available at GitHub: https://github.com/XingyiGuo/Decov-MeWAS.

## Acknowledgements

We thank Dr. Victor Moreno from the Catalan Institute of Oncology (ICO), L’Hospitalet de Llobregat, Barcelona, Spain, for providing DNA methylation and gene expression data from the BarcUVa-Seq study, and for his expert guidance and support throughout this work.

## Figures

**Figure S1.**
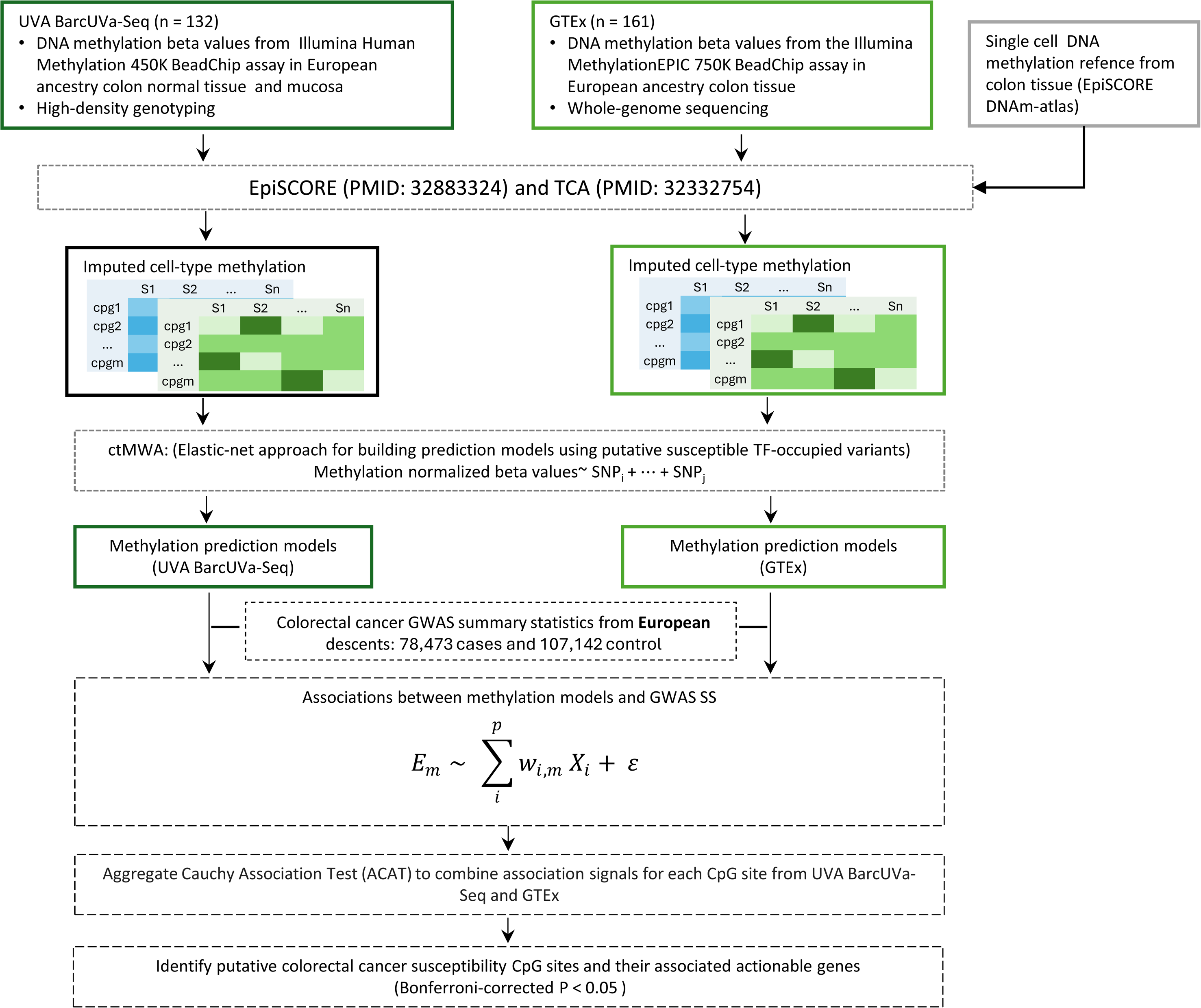
Overview of ctMWAS framework. Workflow for cell-type specific methylation-wide association analysis. Bulk DNAm data were deconvoluted into cell-type specific profiles using single-cell references. Genetic prediction models were constructed and integrated with CRC GWAS summary statistics. Results were combined across datasets via meta-analysis, and CpG sites with Bonferroni-corrected *P* < 0.05 were considered significant.

**Figure S2.**
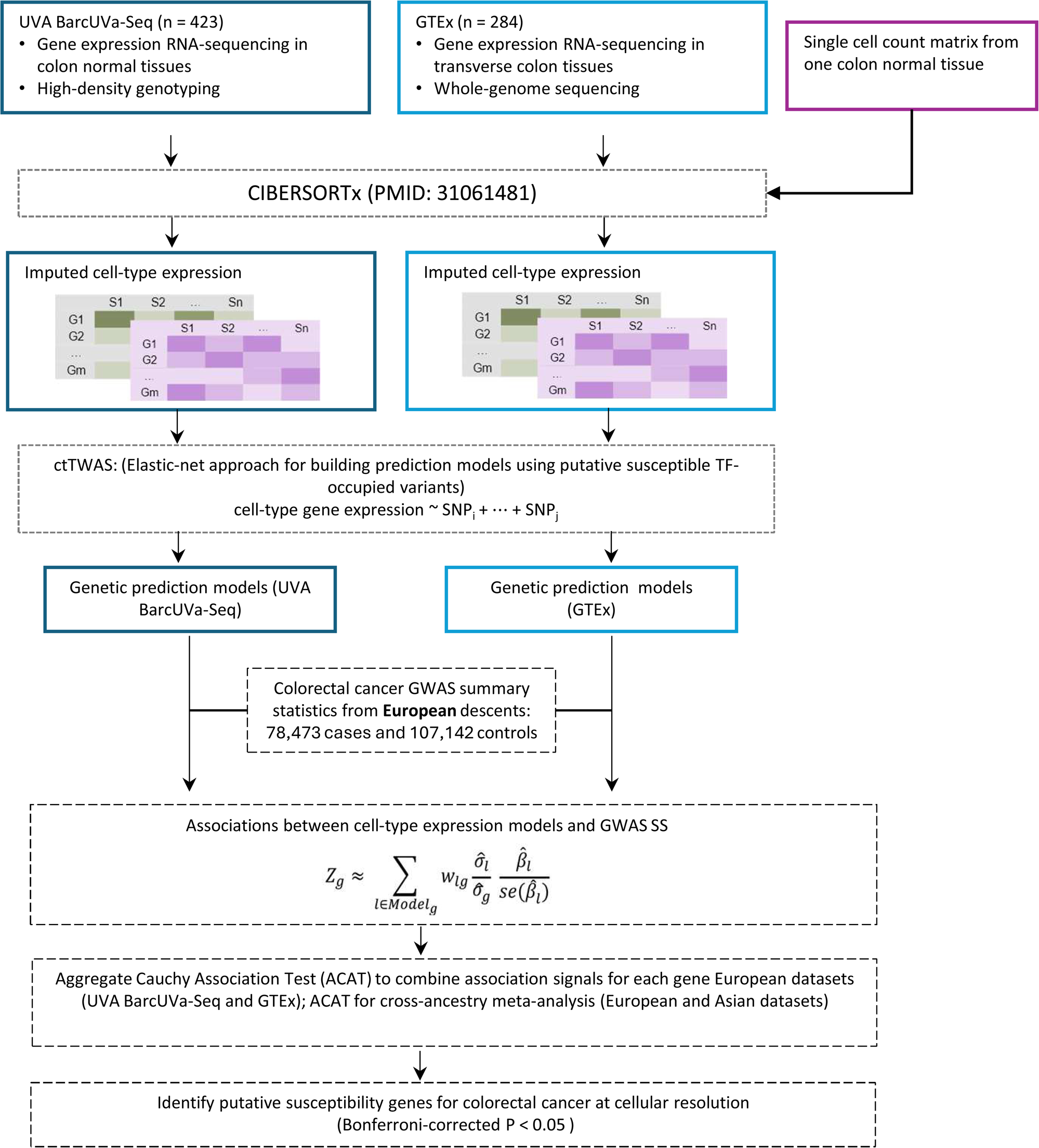
Overview of ctTWAS framework. Workflow for cell-type specific transcriptome-wide association analysis. Bulk RNA-seq data were deconvoluted into cell-type specific expression profiles. Genetic prediction models were integrated with CRC GWAS summary statistics, and results were combined via meta-analysis. Genes with Bonferroni-corrected *P* < 0.05 were considered significant.

**Figure S3.**
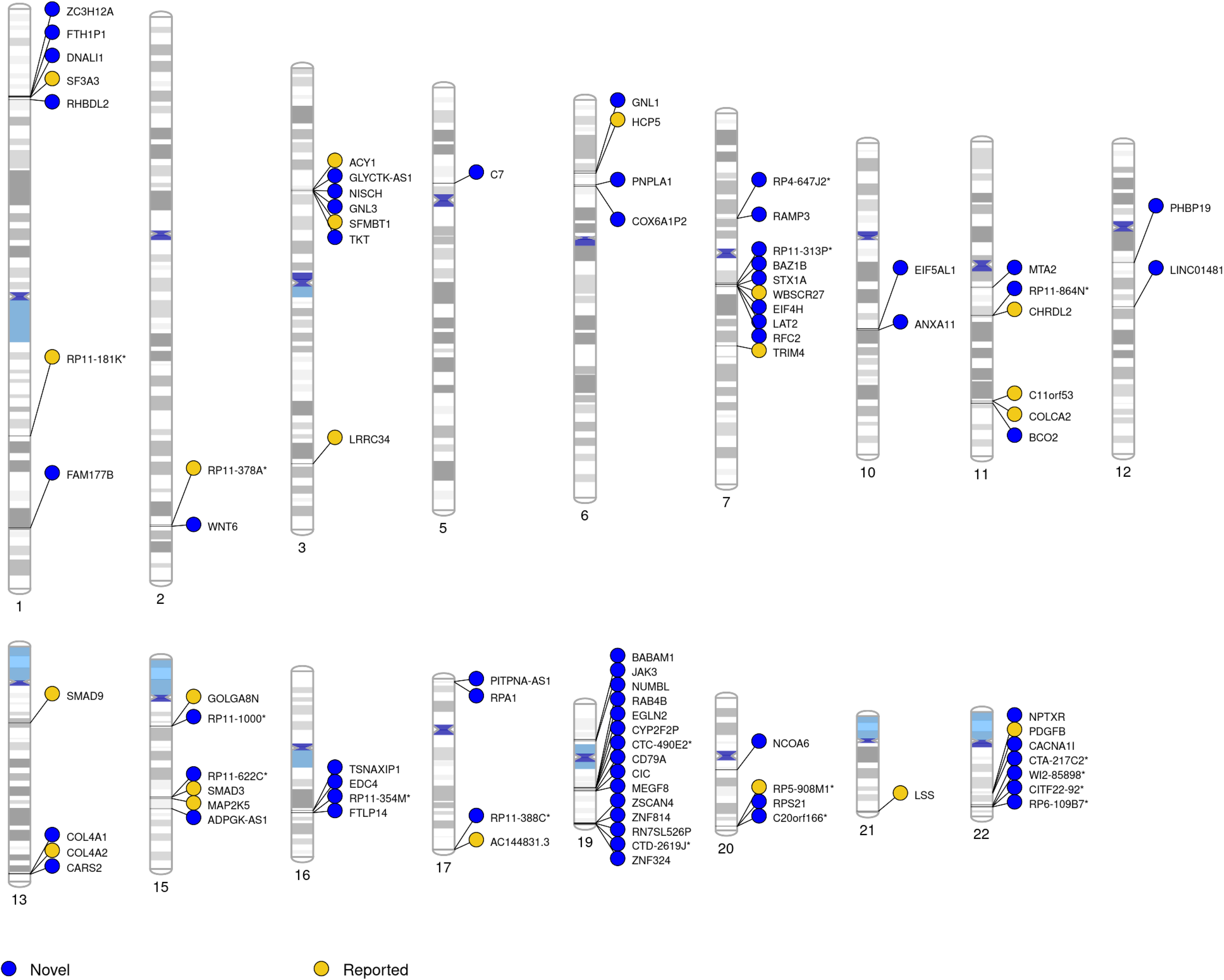
CRC risk genes identified by high confidence GpG sites. Genomic distribution of genes linked to ctMWAS-identified high-confidence CpG sites (PP.H4 > 0.8). Blue circles indicate novel genes identified in this study, and yellow circles represent previously reported genes.

## Supplementary Tables

**Table S1. Estimated cell-type fractions derived from colon DNA methylation data in 132 European-ancestry individuals from the BarcUVa-Seq project.**

**Table S2. Estimated cell-type fractions derived from colon DNA methylation data in 161 European-ancestry individuals from the GTEx project.**

**Table S3. Estimated cell-type fractions derived from colon gene expression data in 423 European-ancestry individuals from the BarcUVa-Seq project.**

**Table S4. Estimated cell-type fractions derived from colon gene expression data in 284 European-ancestry individuals from the GTEx project.**

**Table S5. Significant cell type specific CpG sites identified from ctMWAS analyses across epithelial cell types.**

**Table S6. Significant CpG sites identified from MWAS exhibiting regulatory effects on gene expression based on GTEx data (CpG–gene associations).**

**Table S7. Significant cell type specific genes identified from ctTWAS analyses.**

**Table S8. Multi-omics functional evidence supporting candidate genes identified from ctMWAS and ctTWAS, including TCGA, CPTAC, and DepMap datasets.**

**Table S9. Cell type specific differential expression of CRC risk genes based on single-cell RNA-seq data.**

**Table S10. Pathway enrichment analysis of candidate CRC risk genes identified from ctMWAS and ctTWAS.**

**Table S11. Approved drugs and clinical trial agents targeting candidate CRC risk genes identified from ctMWAS and ctTWAS.**

**Table S12. Concordant candidate genes identified by both ctMWAS and ctTWAS within the same cell type.**

**Table S13. Functional validation of SF3A3 in CRC cell lines (SW480, RKO, HCT116), including cell viability and colony formation assays.**

