## SupplementaryMaterials for "Deconvolution-based cell-type specific DNA methylation-wide and transcriptome-wide association studies identify risk CpG sites and genes associated with colorectal cancer risk"

**Affiliations:**

### Supplementary Methods

#### Construction of cell-type-specific DNA methylation profile

Bulk DNAm processing: DNAm profiles from BarcUVa-Seq were generated using the Illumina HumanMethylation450 array. Standard quality control procedures were applied, including removal of probes with low detection confidence ($P > 0.01$ in > 5% of samples), ambiguous genomic mapping, or nearby SNPs. DNAm data from GTEx (Illumina EPIC array) were processed using comparable pipelines. Detailed experimental procedures have been described previously(1,2).

Estimation of epithelial cell composition: We estimated cell-type proportions using the EpiSCORE framework(3). Proportions were inferred for five major epithelial cells: enteroendocrine, enterocyte, goblet, paneth-like, and progenitor cells. Immune cell types were excluded to maintain consistency with parallel epithelial-focused transcriptomic analyses. Following the EpiSCORE protocol, we summarized DNAm profiles by averaging beta values at CpG sites within ±200 kb of transcription starts sites of cell-type specific marker genes and applied the weighted robust partial correlation (wRPC) algorithm. Cell types with median value of cell fraction larger than 5% among both BarcUVa-Seq and GTEx datasets, were retained for downstream analyses.

Deconvolution of bulk DNAm profiles: We imputed CpG-level, cell type specific DNAm profiles using tensor composition analysis (TCA) (4). TCA tool incorporated key covariates to account for confounding, including age and sex across all datasets, and tissue sites (normal versus mucosa) for BarcUVa-Seq samples. This approach yielded imputed methylation beta values for each CpG across the four epithelial cell types. We further applied quantile normalization, followed by removal of latent confounders using probabilistic estimation of expression residuals (PEER) (5) (20 factors for cohorts with >100 samples, as previously described). The processed DNAm values were then inverse-normal transformed prior to use in downstream prediction model building.

#### Construction of cell-type-specific gene expression profiles

Bulk RNA-seq processing: Bulk RNA-seq data generation, alignment, and initial processing for the BarcUVa-Seq project have been described previously(6,7). Briefly, paired-end RNA sequencing was performed on Illumina platforms, and reads were aligned to the human reference genome (GRCh38 for BarcUVa-Seq and GTEx) using STAR (v2.5.4) (8) following the GTEx Consortium pipeline. Gene expression quantification was conducted using RNA-SeQC (9) based on GENCODE annotations (v26, GRCh38). Following the CIBERSORTx (10) tutorial, we normalized the bulk gene expression count data to Counts Per Million (CPM) for downstream deconvolution analysis.

Cell-type proportion estimation: single-cell RNA sequencing (scRNA-seq) data generated from normal colon tissues from COLON MAP were used to construct epithelial cell-type reference profiles(11). After quality control (filtering low-quality cells and non-epithelial populations), cells were classified into major epithelial subtypes: absorptive cells, crypt top cells, enteroendocrine cells, goblet cells, stem cells, transit amplifying cells, and tuft cells. Each cell contained raw RNA counts spanning around 35,000 genes. We excluded immune and other non-epithelial or low-quality cells and applied quality control filters to retain cells meeting the following criteria: nUMI > 500, nGene > 250, log10GenesPerUMI > 0.8, and mitoRatio < 0.2. The filtered single-cell expression matrix was then processed in CIBERSORTx to construct a cell-type–specific signature matrix. Using the "Impute Cell Fraction" module, cell-type proportions in bulk gene expression data were estimated based on this signature matrix.

Deconvolution of bulk gene expression: After estimating cell-type proportions from the bulk gene expression datasets, we applied the CIBERSORTx "Impute Cell Expression" module to deconvolute bulk gene expression into cell type specific expression profiles. This approach enabled the imputation of cell type–specific gene expression (measured by CPM) for each sample across in BarcUVa-Seq and GTEx datasets. The imputed cell type specific expression values were further converted from CPM to transcripts per million (TPM) within each dataset, followed by quantile normalization and rank-based inverse normal transformation. To account for potential confounding factors, we performed PEER analysis, following GTEx guidelines to determine the appropriate number of factors. Specifically, we used 60 PEER factors for colon tissue in the BarcUVa-Seq and 45 factors for the GTEx transverse colon dataset. The processed expression values were further adjusted for age, sex, and the first five genetic principal components and were then inverse-normal transformed prior to use in downstream prediction model building.

**Genotype data processing**

The process of genotype data from BarcUVa-Seq and GTEx project was described in our previous study(6,7). In general, 400,000 SNPs (Illumina Multi-Ethnic Genotyping Array) from the BarcUVa-Seq project were imputed using TOPMed (Version 2)(12) with European ancestry as reference panel. We then excluded variants with an imputation quality below $R^2 < 0.3$, missingness rate greater than 10%, minor allele frequency (MAF) less than 0.01 (MAF < 0.01), or significant deviations from Hardy-Weinberg equilibrium ($P < 10^{-6}$). In the GTEx project, whole genome sequencing (WGS) was performed on DNA samples from 284 individuals of European ancestry. We downloaded WGS data in the VCF format from dbGaP (phs000424.v8.p2). A comprehensive outline of WGS variant quality control procedures is available in GTEx (1). We removed variants with minor allele frequency (MAF) less than 0.01 (MAF < 0.01), or significant deviations from Hardy-Weinberg equilibrium ($P < 10^{-6}$).

**Mapping ctMWAS-identified CpGs to putative target genes**

To link ctMWAS-identified risk CpG sites to their putative target genes, we evaluated associations between CpG methylation and nearby gene expression (±1 Mb) using matched colon tissue data from individuals of European ancestry. These analyses leveraged 62 overlapping samples with matched DNA methylation (N = 161) and gene expression (N = 284) datasets from the GTEx project. Cell-type–specific methylation and expression profiles were quantile-normalized, adjusted for latent confounders using PEER factors, and inverse-normal transformed prior to analysis (as described in the proceeding sections). For each gene, CpG–gene associations were first assessed using univariate linear regression. For genes associated with multiple CpG sites, we applied multivariate forward selection to identify independent CpG predictors, retaining associations that remained significant in the final model ($P < 0.01$). To prioritize biologically plausible regulatory relationships, we restricted analyses to inverse associations ($\beta < 0$), consistent with the canonical repressive effect of DNA methylation on gene expression.

**Colocalization analyses for significant CpGs and genes**

For CpGs and genes surviving Bonferroni correction, we performed colocalization analyses using the coloc.abf function from the R package coloc (13). We extracted GWAS summary statistics and cell-type-resolved mQTL/eQTL data within ±500 kb windows centered on each significant CpG or gene transcription start site, restricting analyses to loci with ≥ 50 overlapping variants. Applying default priors, we estimated posterior probabilities (PPs) under five competing hypotheses and adopted PP.H4 > 0.8 as the threshold for colocalization, indicating a shared causal variant between the molecular trait and the GWAS signal.

**Inference of oncogenic and tumor-suppressive roles of ctMWAS-identified CpG-associated genes**

We inferred the functional roles of genes by integrating CpG–gene regulatory directionality with ctMWAS association signals. Given that decreased methylation is generally associated with increased gene expression, CpGs with negative ctMWAS Z-scores ($Z < 0$; decreased CRC risk) were interpreted as leading to increased gene expression associated with higher disease risk. Genes linked to such CpGs were therefore classified as putative oncogenes. Conversely, CpGs with positive ctMWAS Z-scores ($Z > 0$; increased CRC risk) were interpreted as leading to reduced gene expression associated with increased disease risk. Genes linked to these CpGs were classified as putative tumor suppressors. This framework provides a mechanistic interpretation of ctMWAS signals by linking genetically regulated DNA methylation to downstream gene expression and their functional roles in CRC susceptibility.

**Characterize potential therapeutic drug candidates**

To identify potential therapeutic drug candidates, we assembled a comprehensive set of drug-target pairs from four drug databases: DrugBank (14), ChEMBL(15), the Therapeutic Target Database (16), and Open Targets (17). DrugBank is a curated database containing over 500,000 drugs and thousands of bioentities (proteins, genes, and other cellular components). ChEMBL provides detailed information on more than 2 million bioactivity assays for bioactive molecules with drug-like properties, covering over 2 million compounds. The Therapeutic Target Database (TTD) offers comprehensive information on thousands of drug targets, including 426 approved, 1014 in clinical trials, 212 in preclinical or patented stages, and 1479 reported in the literature. The Open Targets Platform is a publicly available resource designed to systematically identify and prioritize drug targets. Drugs and their paired genes were comprehensively downloaded from these four databases. We subsequently linked these drugs to our identified disease-susceptibility genes.

**Cell viability assay**

Cell viability was determined using the Alamar Blue (Thermo Fisher) assay. Briefly, SW480, RKO and HCT116 cells were plated at $5 \times 10^3$ cells/well in 96-well plates and reverse-transfected with SF3A3 siRNA (Horizon, ON-TARGETplus Human *SF3A3* (10946) siRNA - SMARTpool, Catalog ID: L-019808-00-0005), a positive control siRNA (Qiagen ,AllStars Hs Cell Death siRNA, catalog no. 10272990), siGENOME Non-Targeting siRNA Pool #1 (Horizon,Catalog No. D-001206-13-05), using RNAiMAX (Life Technologies) according to the manufacturer's instructions. After 96 hours post-transfection, 10 µl of alamarBlue reagent was added to each well (1:10 dilution), incubated at 37°C for 4–6 h, and fluorescence (ex570 nm/em585 nm) was measured on a BioTek Synergy HT plate reader. Percent relative cell viability was calculated as: (mean siSF3A3 value/ mean NC siRNA value) × 100. Three independent experiments were performed for all three cell lines, each repeated in quadruplicate.

**Cell migration and invasion assay**

Cell migration and invasion assays were performed using 24-well plates with 8 µm pore-size chamber inserts (Corning, New York, NY). For the invasion assay, the upper chamber was pre-coated with Corning Cell migration and invasion assays were performed in 24-well plates using inserts with an 8 µm pore size (Millipore) and coated with (invasion assay) or without (migration assay) Corning Matrigel matrix, as described previously (18). SW480, RKO and HCT116 cells were transfected with siSF3A3 or control siRNA. After 16 hours, the cells were detached using trypsin, washed with PBS, and resuspended in serum-free medium. A total of $2 \times 10^4$ cells in 150 µL serum-free medium was seeded into the upper chamber. 500 µL of medium containing 20% FBS was added to the lower chamber as a chemoattractant. After incubation at 37 °C for 36 or 48 hours, cells that had migrated or invaded and adhered to the lower surface of the membrane were fixed in 4% paraformaldehyde for 30 minutes, stained with 0.1% crystal violet, and examined under a light microscope. For quantification, five random visual fields per insert were selected and counted.

**Colony formation assay**

For the colony formation assay, *SF3A3* siRNA-transfected SW480, RKO and HCT116 cells (16 hours post-transfection) were seeded into 6-well plates at a density of 1,000 cells/well, and cultured for 7–14 days. Colonies, defined as those containing ≥ 50 cells, were fixed with methanol, stained with 0.1% (w/v) crystal violet (Sigma-Aldrich), scanned, and counted using ImageJ via batch analysis with a custom plug-in macro. Colony formation efficiency (CFE) was normalized to the negative control (NC) siRNA group and expressed as the percentage (%) of negative control.
